# Data-Driven Biological Subtypes of Parkinson’s Disease

**DOI:** 10.64898/2026.07.28.26359072

**Authors:** Piergiorgio Grillo, Chan Wang, Antonio Pisani, Un Jung Kang, Seyed-Mohammad Fereshtehnejad, Giulietta Maria Riboldi

## Abstract

The heterogenous clinical and pathophysiological presentation of idiopathic Parkinson’s Disease (iPD) suggests the existence of underlying distinct biological subtypes. A better definition of PD subtypes is crucial for the development of target disease modifier approaches. Using data from the Parkinson’s Precision Medicine Initiative (PPMI) cohort, we aimed to identify distinct biological subtypes of iPD through data-driven cluster analysis leveraging currently available biomarker data.

We included n=225 *de novo* iPD subjects. Individuals with known genetic forms of PD were excluded. A total of 22 biomarkers reflecting different biological pathways (neurodegeneration, proteinopathy, neuroinflammation, mitochondrial impairment, and lipid metabolism/autophagy-lysosomal dysfunction) and measured in cerebrospinal fluid (CSF), whole blood, serum, plasma, or urine were selected. Hierarchical clustering was performed using ward.D2 method and Gower dissimilarity. Clinical-demographic characteristics of the identified clusters were compared at baseline and 5-year follow-up.

We identified three clusters: Cluster I (“Mitochondrial-Predominant” subtype); Cluster II (“Cryptic” subtype); Cluster III (“Mixed Pathology” subtype). The “Mitochondrial-Predominant” and “Mixed Pathology” subtypes showed higher levels of CSF mitochondrial DNA deletions and ND1 compared to the “Cryptic” subtype. The “Mixed Pathology” subtype was also characterized by higher levels of serum NfL, CSF p-tau, CSF GFAP, CSF YKL-40, and lower levels of plasma total ceramide, glucosylceramide, and sphingomyelin compared to the other subtypes. In the “Cryptic” subtype no clear dominant biological profile was detected. Clinically, subjects belonging to the “Mixed Pathology” subtype were older and predominantly male. No differences in 5-year clinical progression were found between clusters.

While most of the previous research has performed a biological characterization of the clinical subtypes of PD, with our work we propose a biology-driven clustering in a large cohort if subjects with iPD. We identified three biological subtypes. The “Mitochondrial-Predominant” subtype was characterized by alterations exclusively of biomarkers reflecting mitochondrial dysfunction. The “Mixed Pathology” subtype, in contrast, was characterized by biomarker alterations across all explored pathological pathways. Finally, the “Cryptic” subtype included subjects whose biomarker signature did not indicate predominant alterations in any of the explored pathways. The clusters did not differ in terms of baseline clinical-demographic characteristics, except for age and sex. No difference in mid-term clinical progression was observed between subtypes.

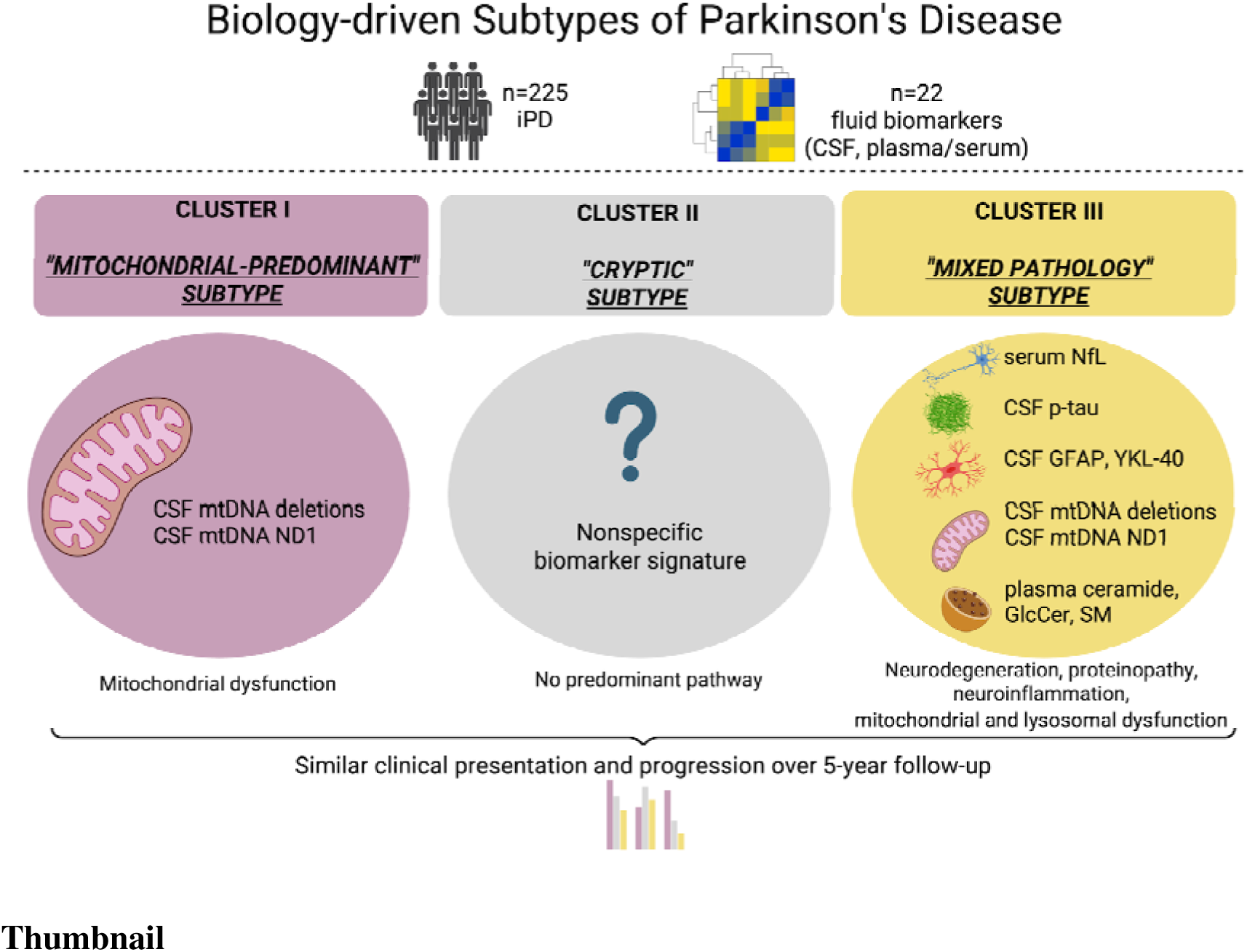

## Introduction

Idiopathic Parkinson’s disease (iPD) is increasingly recognized as a clinically and biologically heterogeneous disorder rather than a single disease entity^1,2^. Clinically, iPD encompasses a broad spectrum of motor and non-motor manifestations, which vary considerably across individuals^2–4^. Likewise, the pathogenesis of iPD is complex and involves multiple pathogenic processes, which are likely represented to different extents across individuals with iPD^5,6^. Among these pathways, the accumulation of α-synuclein aggregates remains central, with cerebrospinal fluid α-synuclein seed amplification assay (CSF-αSyn-SAA) detecting pathological α-synuclein in vivo in approximately 90% of individuals with iPD^7–11^. However, growing evidence indicates that amyloid-β and phosphorylated tau also frequently accumulate in iPD, suggesting that Alzheimer’s Disease (AD) co-pathology is common^12–17^. In addition, insights from monogenic forms of PD support a role for mitochondrial and lysosomal dysfunction in iPD, while evidence from animal models, neuroimaging studies, and laboratory findings from human cohorts suggests the involvement of activated microglia and dysfunctional astrocytes^6,18,19^. These pathogenic processes ultimately converge on the degeneration of several brain regions, including the dopaminergic neurons within the nigrostriatal pathway, which is the key pathological event underlying the characteristic motor symptoms of PD ^5,6^.

The heterogeneity of iPD suggests the existence of distinct disease subtypes that may be identified through data-driven approaches ^1,20^. To date, however, most studies have relied on clinical features to define subtypes, stratifying patients according to their clinical manifestations, whereas biological heterogeneity has received comparatively less attention^3,21^. By contrast, we sought to change this paradigm by identifying subtypes directly from biomarkers reflecting the major pathogenic pathways implicated in iPD. This biology-oriented approach has the potential to better capture the underlying disease heterogeneity and represents an important step toward the development of targeted therapies^20,22,23^.

Within this perspective, we performed a cluster analysis in 225 individuals with iPD from the Parkinson’s Precision Medicine Initiative (PPMI) cohort, using a set of 22 biomarkers measured in CSF, whole blood, serum, plasma, or urine, and reflecting the following biological pathways: neurodegeneration, proteinopathy, neuroinflammation, mitochondrial dysfunction, and lipid metabolism/autophagy-lysosomal dysfunction. Following identification, the resulting biological subtypes were compared in terms of clinical and demographic characteristics, and their clinical progression over a 5-year follow-up period.

## Materials And Methods

The present study was conducted using clinical and biochemical data from PPMI repository, downloaded in June 2025. Complete descriptions of data collected by the PPMI study and information about ethical approval and patient consents can be found at www.ppmi-info.org. Only individuals with an idiopathic form of PD (iPD), testing positive on CSF-αSyn-SAA, were included. Individuals with known genetic forms of PD were excluded. All subjects were drug-naïve, newly diagnosed (within 2 years), here referred to as *de novo* PD. As per PPMI design, diagnosis of PD required the presence of both abnormal dopamine transporter (DaT)-SPECT and two of either resting tremor, bradykinesia, rigidity, or asymmetric resting tremor, or asymmetric bradykinesia ^24^.

All fluid biomarkers available in the PPMI repository as of June 2025 were considered for potential inclusion in this study, except for those derived from omics analyses. Biomarkers measured in CSF, plasma, serum, whole blood, and urine, reflecting different pathological processes, were considered together with amplification parameters from CSF-αSyn-SAA (Figure 1).

**Figure 1.**
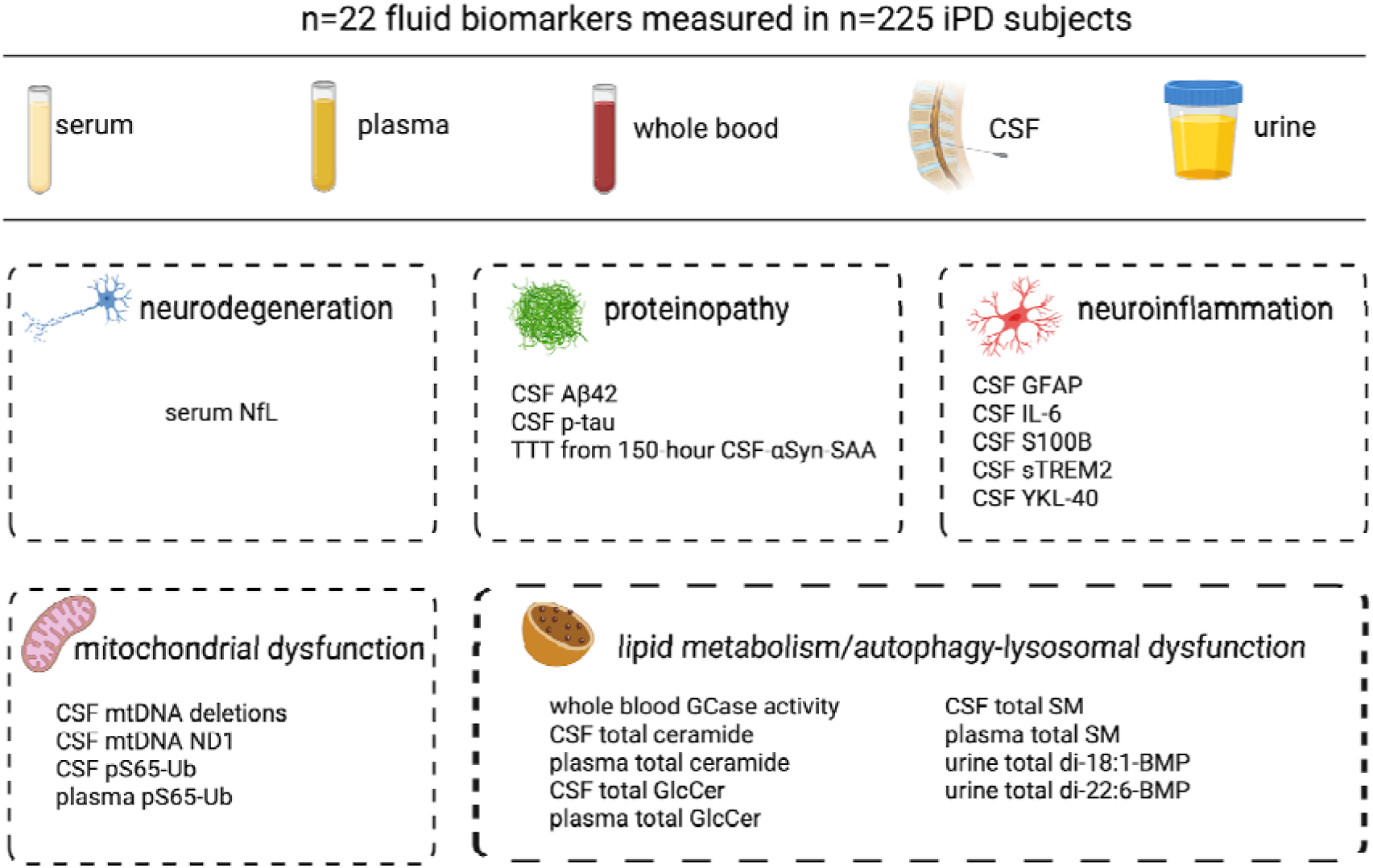
Fluid biomarkers included in the cluster analysis. Illustration of the fluid biomarkers selected for the cluster analysis, categorized according to the pathological pathways they represent. Abbreviations: n, number; CSF, cerebrospinal fluid; serum NfL, serum neurofilament light chain; CSF Aβ42, CSF amyloid-beta 42; CSF p-tau, CSF phosphorylated tau; TTT from 150-hour CSF-αSyn-SAA, Time to Threshold from 150-hour cerebrospinal fluid α-synuclein seed amplification assay; CSF GFAP, CSF glial fibrillary acidic protein; CSF IL-6, CSF interleukin-6; CSF sTREM2, CSF soluble triggering receptor expressed on myeloid cells 2; CSF mtDNA deletions, CSF mitochondrial DNA deletions; CSF mtDNA ND1, CSF mitochondrial DNA NADH dehydrogenase subunit 1; CSF and plasma pS65-Ub, CSF and plasma phosphorylated serine-65 ubiquitin; whole blood GCase activity, whole blood glucocerebrosidase activity; CSF and plasma total GlcCer, CSF and plasma total glucosylceramide; CSF and plasma total SM, CSF and plasma total sphingomyelin; urine total di-18:1-BMPand total di-22:6-BMP, urine total di-18:1- and total di-22:6-bis(monoacylglycerol)phosphate. Created with BioRender. Pisani, A. (2026) https://BioRender.com/hqnddr1

### Selection Process of Fluid Biomarkers for Cluster Analysis

The selection of fluid biomarkers for cluster analysis followed a three-step process:

-First, fluid biomarkers available for less than n=100 individuals with iPD were excluded to maximize the statistical power.

-The selected biomarkers were not necessarily measured in the same participants. To minimize missing data for the cluster analysis, we selected the largest possible set of biomarkers with the highest degree of overlap across subjects. As a part of this process, αSyn seed amplification assay (SAA) parameters derived from the 150-hour CSF-αSyn-SAA were selected over those derived from the 24-hour CSF-αSyn-SAA. Description of the two different assays and corresponding amplification parameters has been reported previously^10,11,25,26^.

-Third, when two or more biomarkers in related biological pathways were highly correlated - an absolute Pearson correlation coefficient>0.8 - only the biomarker with the highest clinical relevance was retained (Supplementary Figure 1). Accordingly, CSF phosphorylated tau (p-tau) was selected over CSF total tau (t-tau) ^12,13,17^; CSF mitochondrial DNA (mtDNA) NADH dehydrogenase subunit 1 (ND1) was selected over CSF mtDNA NADH dehydrogenase subunit 4 (ND4)^27–30^; CSF and plasma total glucosylceramide (GlcCer) were selected over CSF and plasma total lactosylceramide (GL2) ^31–33^; urine total di-22:6-bis(monoacylglycerol)phosphate (BMP) was selected over urine 2,2’ di-22:6-BMP ^34^. Among the 150-hour CSF-αSyn-SAA amplification parameters, time to threshold (TTT) was selected over time to reach 50% of Fmax (T50), maximum fluorescence (Fmax), slope, and area under the curve (AUC), as previous studies have shown that this parameter correlates more consistently with clinical features^7,11,35^.

At the end of the third and last step of selection, a total of 22 out of more than 100 fluid biomarkers were retained for the final analysis in a sample of 225 individuals (Figure 1).

### Fluid Biomarkers Included in the Cluster Analysis

Cluster analysis was performed on a sample of 225 individuals with iPD, for whom 22 continuous fluid biomarkers retained through the selection process were available (Figure 1). All biomarkers were obtained at baseline, in the drug-naïve stage. Biomarkers were grouped into five categories reflecting distinct pathological processes based on known biological function and previous literature:

-Markers of neurodegeneration ^36,37^: serum neurofilament light chain (NfL)

-Markers of proteinopathy ^7,12,35^: CSF amyloid-beta 42 (Aβ42), CSF phosphorylated tau (p-tau), and Time to Threshold (TTT) from 150-hour CSF-αSyn-SAA.

-Markers of neuroinflammation^37,38^: CSF glial fibrillary acidic protein (GFAP), CSF interleukin-6 (IL-6), CSF S100B, CSF soluble triggering receptor expressed on myeloid cells 2 (sTREM2), and CSF YKL-40.

-Markers of mitochondrial dysfunction^27–30^: CSF mitochondrial DNA (mtDNA) deletions, CSF mitochondrial DNA (mtDNA) NADH dehydrogenase subunit 1 (ND1), and CSF and plasma phosphorylated serine-65 ubiquitin (pS65-Ub)

-Markers of lipid metabolism/autophagy-lysosomal dysfunction^31–34,39,40^: whole blood glucocerebrosidase (GCase) activity, CSF and plasma total ceramide, CSF and plasma total glucosylceramide (GlcCer), and CSF and plasma total sphingomyelin (SM), as well as urine total di-18:1- and di-22:6-bis(monoacylglycerol)phosphate (BMP)

### Clinical and Demographic Characterization of Clusters at Baseline and 5-Year Follow-Up

After cluster identification, the following variables were used to characterize each cluster at baseline and at the 5-year follow-up:

-Demographic parameters: age, sex, race, education level, body mass index;

-Clinical parameters: family history of PD, disease duration, Movement Disorder Society-Unified PD Rating scale (MDS-UPDRS) part I, II, III, IV and total score, Hoehn and Yahr scale (H&Y), levodopa equivalent daily dose (LEDD), Montreal cognitive assessment (MoCA), Scales for Outcomes in PD-Autonomic dysfunction (SCOPA-AUT), RBD Screening Questionnaire (RBDSQ), University of Pennsylvania Smell Identification Test (UPSIT), State-Trait Anxiety Inventory (STAI). Clinical differences between clusters were further assessed by comparing the overlap of clinical subtypes defined according to two established classification systems. The first classification system distinguishes tremor-dominant PD from postural instability and gait disorder (PIGD) PD based exclusively on motor symptom features^21^. The second classification system identifies mild motor-predominant (MMP), intermediate (IM), and diffuse malignant (DM) PD based on a composite score incorporating motor, sleep, cognitive, and dysautonomia features^3^. The occurrence of the following clinical milestones, as defined previously, was also assessed at 5-year follow-up: activities of daily living impairment, autonomic dysfunction, cognitive impairment, functional dependence, motor complications, and walking and balance impairment^41^.

### Statistical Analysis

Biomarker values below the limit of detection were replaced with the corresponding detection-limit values and log2-transformed, when appropriate, to reduce skewness and stabilize variance. Hierarchical clustering was performed using ward.D2 method based on Gower dissimilarity. Missing data were retained, and Gower dissimilarities were computed using the available observed measurements for each pair of participants. The optimal number of clusters was determined by maximizing the average silhouette width across candidate solutions (k = 2–8).

To characterize cluster profiles, we calculated an effect measure defined as the signed standardized difference between each cluster-specific mean and the overall mean for each variable, computed as (cluster mean − overall mean) / overall standard deviation. Positive values indicate that the cluster mean was above the overall average, whereas negative values indicate that it was below the overall average; the magnitude reflects the distance of the cluster mean from the overall mean in standard deviation units.

To evaluate associations between cluster membership and clinical or demographic characteristics at baseline and 5-year follow-up, categorical variables were compared across clusters using chi-square tests, and continuous variables were compared using analysis of variance (ANOVA) followed by Tukey-adjusted post hoc pairwise comparisons. Associations between cluster membership and clinical subtype at baseline and year 5 were assessed using chi-square tests or Fisher’s exact tests, as appropriate. All statistical analyses were performed using R (R Core Team, R Foundation of Statistical Computing) version 4.4.3.

## Results

Demographic and clinical features of the study cohort at baseline (n=225) are reported in Table 1.

**Table 1.** Demographic and clinical parameters of the study cohort at baseline. Continuous variables are presented as mean ± standard deviation, and categorical variables as number, frequency.; Abbreviations: n, number; BMI, body mass index; LEDD, levodopa equivalent daily dose; MDS-UPDRS-part I, II, III, IV and total score, Movement Disorder Society-Unified PD Rating scale part part I, II, III, IV and total score; HY, Hoehn and Yahr scale; MoCA, Montreal cognitive assessment; SCOPA-AUT, Scales for Outcomes in PD-Autonomic dysfunction; RBDSQ, RBD Screening Questionnaire; UPSIT, University of Pennsylvania Smell Identification Test; STAI, State-Trait Anxiety Inventory; PIGD, Postural Instability and Gait Difficulty.

| Study Cohort<br>N=225 |  |
| --- | --- |
| Demographic Parameters |  |
| Age (years) | 61.6 ± 9.1 |
| Sex (male, %) | 144, 64.0% |
| Race (white, %) | 208, 92.4% |
| Education (years) | 15.9 ± 2.5 |
| BMI | 27.1 ± 4.6 |
| Clinical Parameters |  |
| Family history of PD (yes, %) | 59, 26.2% |
| Duration (years) | 0.6 ± 0.6 |
| LEDD (mg/day) | 0 |
| MDS-UPDRS-part I | 5.6 ± 3.9 |
| MDS-UPDRS-part II | 5.9 ± 4.2 |
| MDS-UPDRS-part III<br>in OFF state | 21.5 ± 9.2 |
| MDS-UPDRS-part IV | 0 |
| MDS-UPDRS total score | 33.1 ± 13.2 |
| HY in OFF state | 1.5 ± 0.5 |
| MoCA | 27.1 ± 2.4 |
| SCOPA-AUT | 9.6 ± 6.4 |
| RBDSQ | 4.0 ± 2.7 |
| UPSIT | 21.5 ± 7.8 |
| STAI | 65.2 ± 18.1 |
| PIGD<br>phenotype (yes, %) | 55 ± 24.4% |
| Diffuse Malignant<br>phenotype (yes, %) | 27, 12.2% |

### Identification of Biological Subtypes by Cluster Analysis

The cluster analysis identified three clusters (Supplementary Figure 2 and 3, Figure 2 and 3, Table 2).

**Figure 2.**
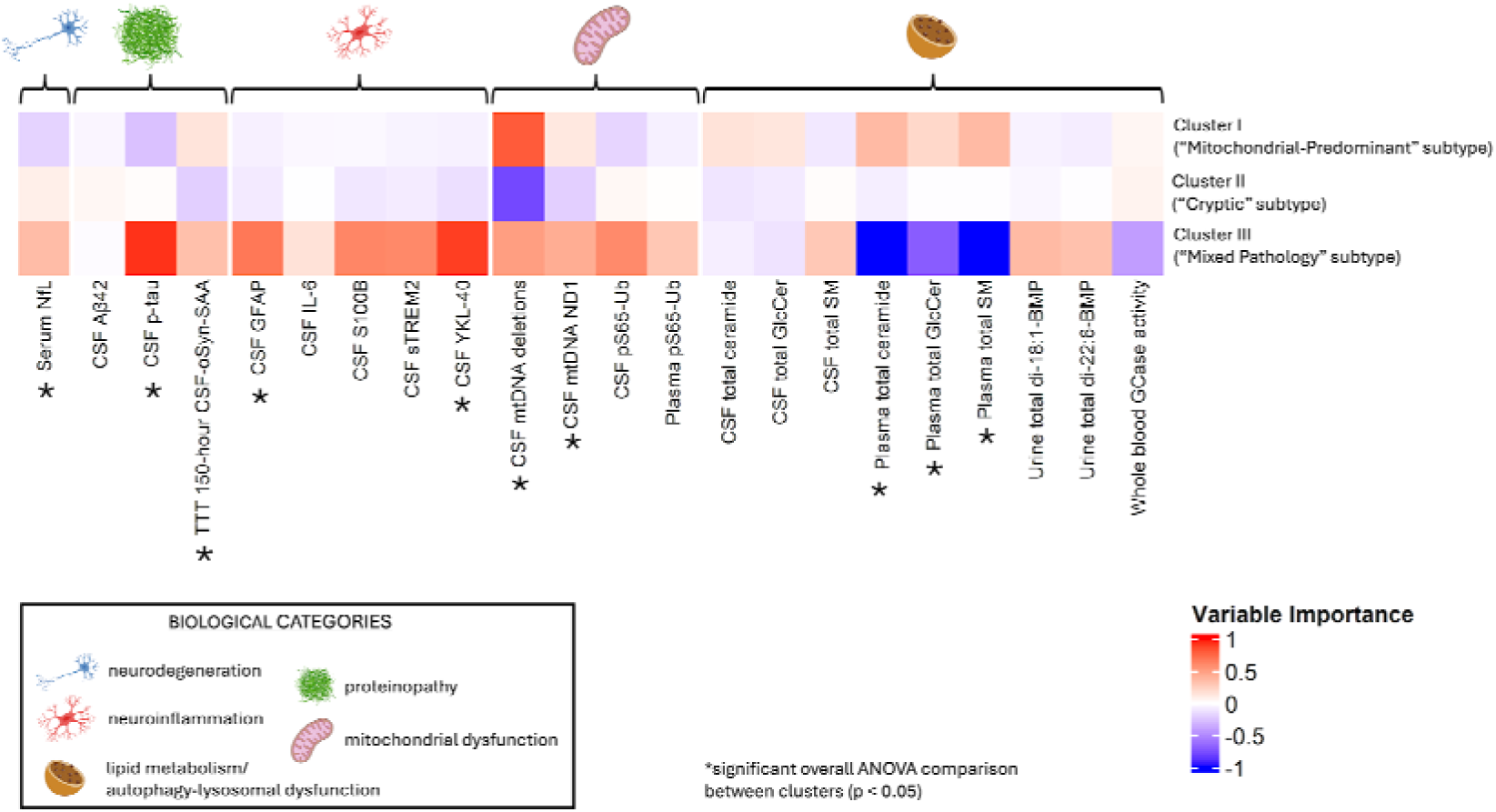
Heatmap showing normalized variable importance across clusters. Values represent signed standardized differences for each feature, calculated as the difference between the cluster-specific mean and the overall mean, scaled by the overall standard deviation. Positive values indicate that the cluster mean is above the overall average for that variable, whereas negative values indicate that the cluster mean is below the overall average. The magnitude reflects the distance of the cluster mean from the overall mean in standard deviation units. Variables are grouped by biological category, indicated by distinct icons. Cluster I was renamed as the “Mitochondrial-Predominant” subtype; Cluster II was renamed as the “Cryptic” subtype; Cluster III was renamed as the “Mixed Pathology” subtype *Indicates a statistically significant overall ANOVA comparison between clusters (p < 0.05).

**Figure 3.**
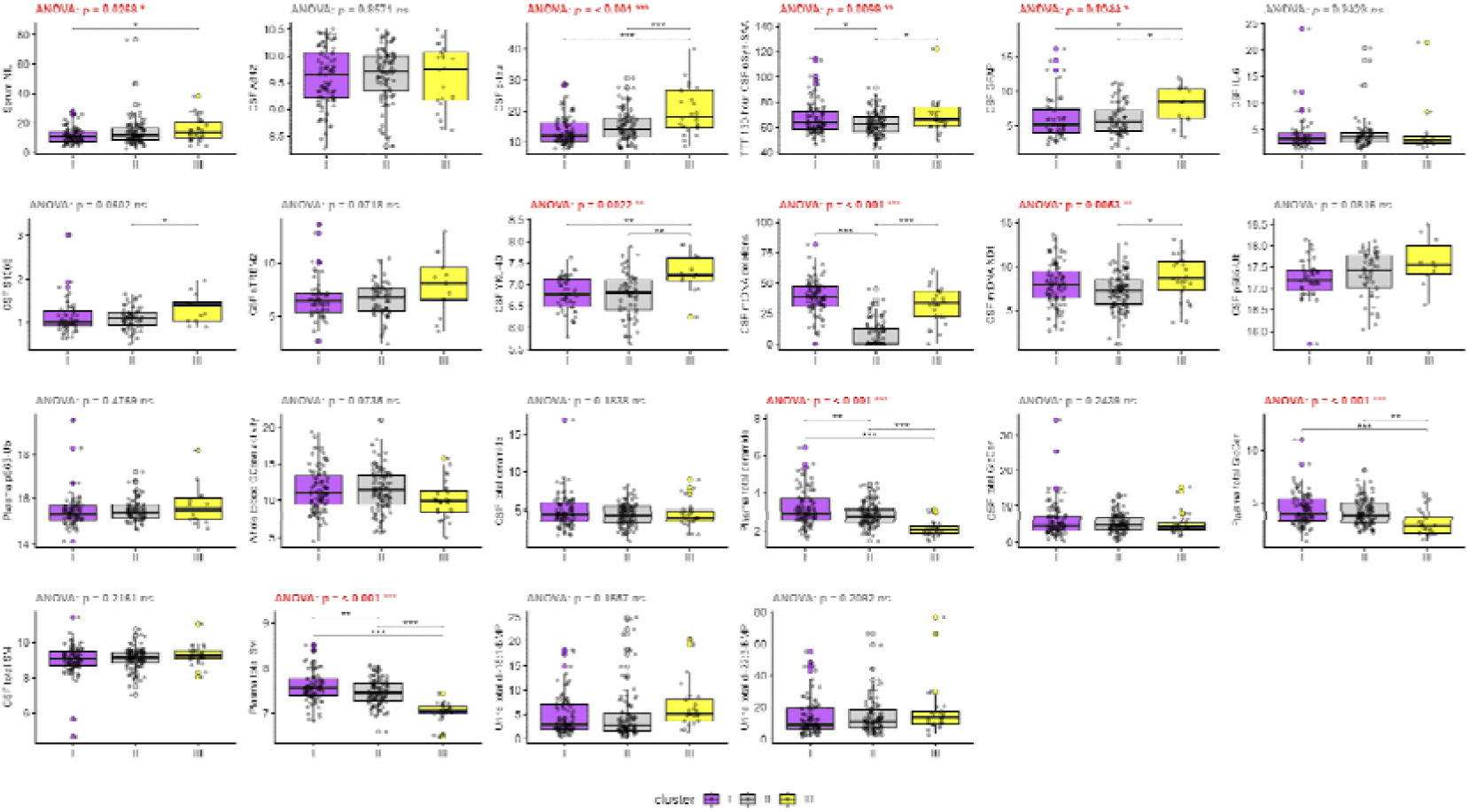
Distribution of biomarkers across the three data-driven clusters. Each panel shows one biomarker, with boxplots summarizing values within each cluster and overlaid points representing individual subjects. The center line indicates the median, the box bounds indicate the first and third quartiles, and the whiskers extend to the minimum and maximum values. Overall differences across clusters were assessed by analysis of variance (ANOVA), with the corresponding p-value shown at the top of each panel. When the overall ANOVA was significant, Tukey-adjusted post hoc pairwise comparisons were performed and are indicated by brackets. Statistical significance is denoted as * P<0.05, ** P<0.01, and *** P<0.001; ns, not significant. Colors indicate cluster membership (purple, Cluster I, “Mitochondrial-Predominant” subtype; grey, Cluster II, “Cryptic” subtype; yellow, Cluster III, “Mixed Pathology” subtype). Abbreviations: CSF, cerebrospinal fluid; serum NfL, serum neurofilament light chain; CSF Aβ42, CSF amyloid-beta 42; CSF p-tau, CSF phosphorylated tau; TTT from 150-hour CSF-αSyn-SAA, Time to Threshold from 150-hour cerebrospinal fluid α-synuclein seed amplification assay; CSF GFAP, CSF glial fibrillary acidic protein; CSF IL-6, CSF interleukin-6; CSF sTREM2, CSF soluble triggering receptor expressed on myeloid cells 2; CSF mtDNA deletions, CSF mitochondrial DNA deletions; CSF mtDNA ND1, CSF mitochondrial DNA NADH dehydrogenase subunit 1; CSF and plasma pS65-Ub, CSF and plasma phosphorylated serine-65 ubiquitin; whole blood GCase activity, whole blood glucocerebrosidase activity; CSF and plasma total GlcCer, CSF and plasma total glucosylceramide; CSF and plasma total SM, CSF and plasma total sphingomyelin; urine total di-18:1-BMPand total di-22:6-BMP, urine total di-18:1- and total di-22:6-bis(monoacylglycerol)phosphate.

**Table 2.** Comparison of fluid biomarkers across clusters. Values are given in mean ± standard deviation. For overall *p*-value, statistical significance is marked in bold. For *Post Hoc* pairwise comparisons, only statistically significant comparisons are reported.; Abbreviations: n, number; serum NfL, serum neurofilament light chain; CSF Aβ42, CSF amyloid-beta 42; CSF p-tau, CSF phosphorylated tau; TTT from 150-hour CSF-αSyn-SAA, Time to Threshold from 150-hour cerebrospinal fluid α-synuclein seed amplification assay; CSF GFAP, CSF glial fibrillary acidic protein; CSF IL-6, CSF interleukin-6; CSF sTREM2, CSF soluble triggering receptor expressed on myeloid cells 2; CSF mtDNA deletions, CSF mitochondrial DNA deletions; CSF mtDNA ND1, CSF mitochondrial DNA NADH dehydrogenase subunit 1; CSF and plasma pS65-Ub, CSF and plasma phosphorylated serine-65 ubiquitin; whole blood GCase activity, whole blood glucocerebrosidase activity; CSF and plasma total GlcCer, CSF and plasma total glucosylceramide; CSF and plasma total SM, CSF and plasma total sphingomyelin; urine total di-18:1-BMPand total di-22:6-BMP, urine total di-18:1- and total di-22:6-bis(monoacylglycerol)phosphate

|  | Cluster I<br>or<br>“Mitochondrial-Predominant”<br>subtype<br>N=94 | Cluster II<br>or<br>“Cryptic”<br>subtype<br>N=105 | Cluster III<br>Or<br>“Mixed<br>Pathology”<br>subtype<br>N=26 | Overall<br>*p*-value | *Post Hoc*<br>pairwise<br>comparison |
| --- | --- | --- | --- | --- | --- |
| **Markers of neurodegeneration** |  |  |  |  |  |
| serum NfL | 11.6 ± 5.2 | 13.7 ± 9.2 | 15.7 ± 8.3 | **0.0268** | **I versus III** |
| **Markers of proteinopathy** |  |  |  |  |  |
| CSF Aβ42 | 9.6 ± 0.5 | 9.6 ± 0.5 | 9.6 ± 0.6 | 0.8571 | - |
| CSF p-tau | 13.6 ± 4.5 | 15.2 ± 4.9 | 20.2 ± 7.7 | **<0.001** | **I versus III**<br>**II versus III** |
| TTT<br>From 150-hour<br>CSF-αSyn-SAA | 67.1 ± 13.0 | 63.1 ± 9.1 | 69.3 ± 13.7 | **0.0098** | **I versus II**<br>**II versus III** |
| **Markers of neuroinflammation** |  |  |  |  |  |
| CSF GFAP | 6.1 ± 3.1 | 6.0 ± 2.3 | 8.1 ± 2.7 | **0.0344** | **I versus III**<br>**II versus III** |
| CSF IL-6 | 4.1 ± 3.4 | 4.2 ± 3.3 | 4.8 ± 5.3 | 0.8428 | - |
| CSF S100B | 1.1 ± 0.4 | 1.1 ± 0.2 | 1.3 ± 0.3 | 0.0602 | - |
| CSF sTREM2 | 6.6 ± 2.0 | 6.6 ± 1.7 | 7.9 ± 2.8 | 0.0718 | - |
| CSF YKL-40 | 6.8 ± 0.4 | 6.8 ± 0.5 | 7.3 ± 0.5 | **0.0022** | **I versus III**<br>**II versus III** |
| **Markers of mitochondrial dysfunction** |  |  |  |  |  |
| CSF mtDNA deletions | 38.4 ± 14.3 | 7.1 ± 10.3 | 32.5 ± 15.4 | **<0.001** | **I versus II**<br>**II versus III** |
| CSF mtDNA ND1 | 7.9 ± 2.4 | 7.2 ± 2.0 | 8.6 ± 2.5 | **0.0053** | **II versus III** |
| CSF pS65-Ub | 17.2 ± 0.4 | 17.3 ± 0.5 | 17.6 ± 0.6 | 0.0816 | - |
| plasma pS65-Ub | 15.5 ± 0.8 | 15.5 ± 0.5 | 15.7 ± 0.9 | 0.4769 | - |
| **Markers of lipid metabolism and autophagy-lysosomal dysfunction** |  |  |  |  |  |
| whole blood<br>GCase activity | 11.5 ± 2.9 | 11.5 ± 2.9 | 10.1 ± 2.5 | 0.0735 | - |
| CSF total ceramide | 4.8 ± 2.1 | 4.4 ± 1.4 | 4.5 ± 1.7 | 0.1838 | - |
| plasma total ceramide | 3.2 ± 0.9 | 2.9 ± 0.6 | 2.1 ± 0.4 | **<0.001** | **I versus II**<br>**I versus III**<br>**II versus III** |
| CSF total GlcCer | 6.2 ± 4.9 | 5.4 ± 2.6 | 5.3 ± 3.2 | 0.2439 | - |
| plasma total GlcCer | 4.5 ± 1.5 | 4.2 ± 1.2 | 3.2 ± 1.2 | **<0.001** | **I versus III**<br>**II versus III** |
| CSF total SM | 9.0 ± 0.8 | 9.1 ± 0.6 | 9.3 ± 0.5 | 0.2161 | - |
| plasma total SM | 7.6 ± 0.3 | 7.5 ± 0.3 | 7.0 ± 0.2 | **<0.001** | **I versus II**<br>**I versus III**<br>**II versus III** |
| urine total di-18:1-BMP | 4.7 ± 4.0 | 4.8 ± 5.3 | 6.7 ± 4.9 | 0.1657 | - |
| urine total di-22:6-BMP | 13.6 ± 11.3 | 14.4 ± 11.3 | 18.4 ± 17.3 | 0.2092 | - |

-Cluster I, including n=94 subjects, was characterized by predominant differences in markers of mitochondrial dysfunction. Specifically, Cluster I exhibited significantly higher levels of CSF mtDNA deletions and a trend toward higher levels of CSF mtDNA ND1 compared with Cluster II (CSF mtDNA deletions, overall ANOVA *p*<0.001, Cluster I vs Cluster II, 38.4±14.3 vs 7.1±10.3, p<0.001; CSF mtDNA ND1, overall ANOVA *p*=0.0053, Cluster I vs Cluster II, 7.9±2.4 vs 7.2±2.0, *p*=0.061). Although Cluster III also exhibited significantly higher levels of CSF mtDNA deletions and CSF mtDNA ND1 than Cluster II, these changes occurred alongside alterations in biomarkers representing several other pathogenic pathways. (CSF mtDNA deletions, overall ANOVA *p*<0.001, Cluster III vs Cluster II, 32.5±15.4 vs 7.1±10.3, *p*<0.001; CSF mtDNA ND1, overall ANOVA *p*=0.0053, Cluster III vs Cluster II, 8.6±2.5 vs 7.2±2.0, *p*=0.010). Based on these findings, Cluster I was labeled the “Mitochondrial-Predominant” subtype (Supplementary Figure 3, Figure 2 and 3, Table 2).

-Cluster II, including n=105 subjects, was not characterized by any specific biomarker signature. Notably, it was the only cluster showing lower levels of CSF mtDNA deletions and CSF mtDNA ND1 compared with Cluster I, Cluster III, or both, suggesting a relative absence of the mitochondrial alterations hypothesized for the other two clusters. Cluster II also exhibited significantly shorter CSF αSyn-SAA TTT values than Clusters I and III, although this difference resulted in only subtle differences in the relative biomarker values displayed in the heatmap (overall ANOVA *p*=0.0098; Cluster II vs Cluster I, 63.1±9.1 vs 67.1±13.0, *p*=0.037; Cluster II vs Cluster III, 63.1±9.1 vs 69.3±13.7, *p*=0.036). Based on these results, Cluster II was renamed as the “Cryptic” subtype (Supplementary Figure 3, Figure 2 and 3, Table 2).

-Cluster III, eventually, including n=26 subjects, was characterized by alteration across all the pathological pathways explored with fluid biomarkers. In particular, Cluster III showed higher levels of serum NfL, CSF p-tau, CSF GFAP, CSF YKL-40, CSF mtDNA deletions, CSF mtDNA ND1 and lower levels of plasma total ceramide, GlcCer, and SM compared to Cluster I, Cluster II, or both (Table 2). Although statistical significance was not reached, Cluster III also tended to exhibit the highest CSF sTREM2 and S100B levels, and the lowest whole-blood GCase activity among the three clusters (CSF sTREM2: Cluster III 7.9±2.8, Cluster I 6.6±2.0, Cluster II 6.6±1.7, overall ANOVA *p*=0.0718; CSF S100B: Cluster III 1.3±0.3, Cluster I 1.1±0.4, Cluster II 1.1±0.2, overall ANOVA *p*=0.0602; whole-blood GCase activity: Cluster III 10.1±2.5, Cluster I 11.5±2.9, Cluster II 11.5±2.9, overall ANOVA p=0.0735). This suggested the coexistence of prominent neurodegenerative, neuroinflammatory, mitochondrial and lysosomal dysfunction in this cluster. Accordingly, Cluster III was renamed as the “Mixed Pathology” Subtype (Supplementary Figure 3, Figure 2 and 3, Table 2).

### Demographic and Clinical Characterization of Cluster-Derived Biological Subtypes at Baseline

The characterization of cluster-derived biological subtypes at baseline revealed differences only in demographic features. Specifically, individuals belonging to Cluster III (the “Mixed Pathology” subtype) were predominantly male compared to those in the other two clusters, namely Cluster I (the “Mitochondrial-Predominant” subtype) and Cluster II (the “Cryptic” subtype), and older than those in Cluster I (the “Mitochondrial-Predominant” subtype) (male distribution: overall p=0.0154, Cluster III vs Cluster I 88.5% vs 63.8% p=0.0251, Cluster III vs Cluster II 88.5% vs 58.1% p=0.0094; age: overall p=0.0090, Cluster III vs Cluster I 65.9±7.9 vs 59.9±8.8 p=0.0078). No differences were observed in motor or non-motor clinical features. The distribution of clinical subtypes, determined at baseline using the TD/PIGD and MMP/IM/DM classifications, was similar across cluster-derived biological subtypes (Table 3, Figure 4)

**Figure 4.**
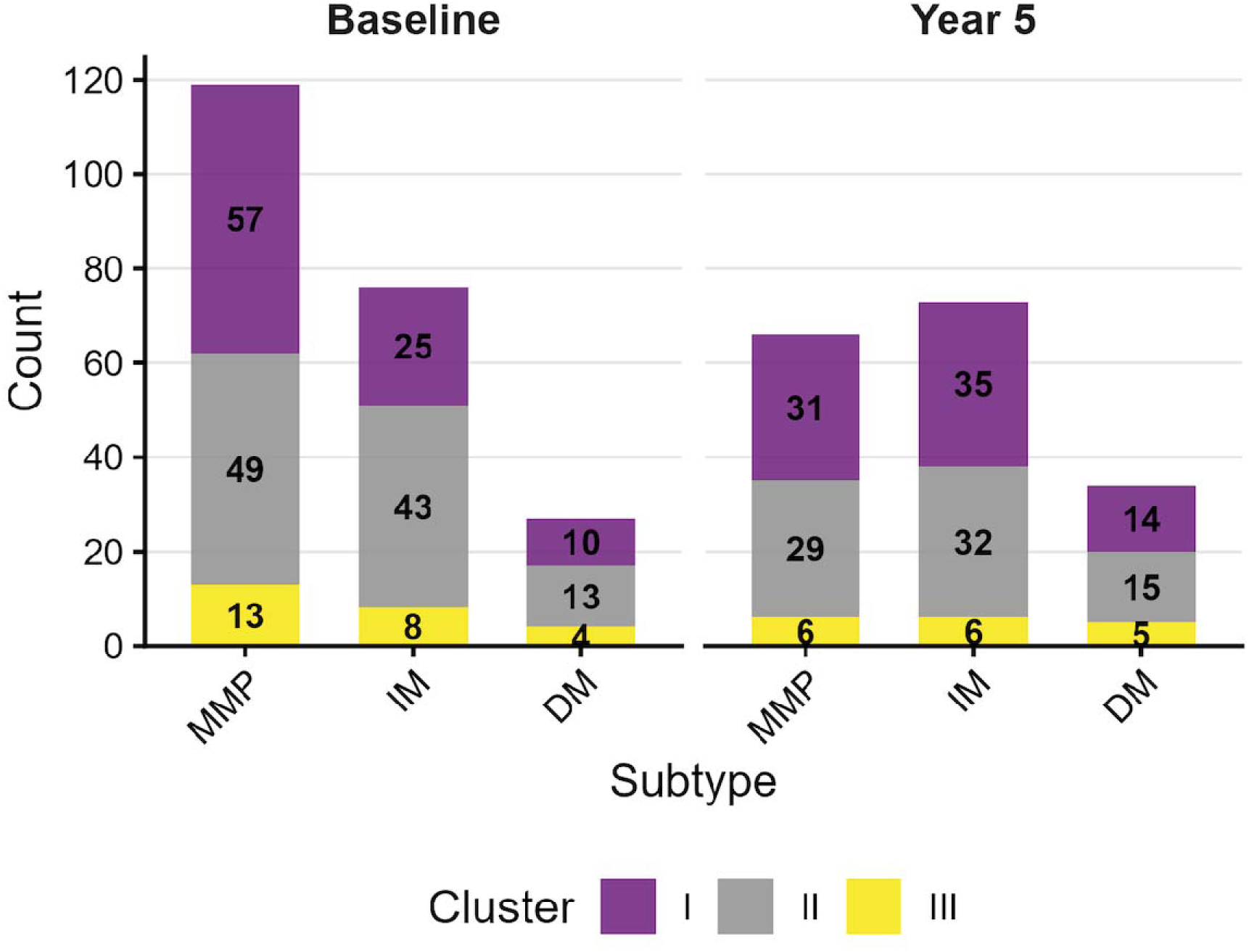
Distribution of biological subtypes across clinical subtypes at baseline and 5-year follow up. Stacked bar plots show the number of participants in each clinical subtype (Mild Motor Predominant, MMP; Intermediate, IM; Diffuse Malignant, DM), stratified by biological subtype membership (purple, Cluster I, “Mitochondrial-Predominant” subtype; grey, Cluster II, “Cryptic” subtype; yellow, Cluster III, “Mixed Pathology” subtype). Numbers within bars indicate participant counts for biological subtype within each clinical subtype.

**Table 3.** Comparison of demographic and clinical parameters across clusters at baseline. Continuous variables are presented as mean ± standard deviation, and categorical variables as number, frequency. For overall *p*-value, statistical significance is marked in bold. For *Post Hoc* pairwise comparisons, only statistically significant comparisons are reported.; Abbreviations: n, number; BMI, body mass index; LEDD, levodopa equivalent daily dose; MDS-UPDRS-part I, II, III, IV and total score, Movement Disorder Society-Unified PD Rating scale part part I, II, III, IV and total score; HY, Hoehn and Yahr scale; MoCA, Montreal cognitive assessment; SCOPA-AUT, Scales for Outcomes in PD-Autonomic dysfunction; RBDSQ, RBD Screening Questionnaire; UPSIT, University of Pennsylvania Smell Identification Test; STAI, State-Trait Anxiety Inventory; PIGD, Postural Instability and Gait Difficulty.

|  | **Cluster I**<br>**or**<br>**“Mitochondrial-**<br>**Predominant”**<br>**subtype**<br>**N=94** | **Cluster II**<br>**or**<br>**“Cryptic”**<br>**subtype**<br>**N=105** | **Cluster III**<br>**Or**<br>**“Mixed**<br>**Pathology”**<br>**subtype**<br>**N=26** | **Overall**<br>**p-value** | **Post Hoc**<br>**pairwise**<br>**comparison** |
| --- | --- | --- | --- | --- | --- |
| **Demographic Parameters** |  |  |  |  |  |
| Age (years) | 59.9 ± 8.8 | 62.1 ± 9.4 | 65.9 ± 7.9 | **0.0090** | **I versus III** |
| Sex (male, %) | 60, 63.8% | 61, 58.1% | 23, 88.5% | **0.0154** | **I versus III**<br>**II versus III** |
| Race (white, %) | 87, 93.5% | 96, 91.4% | 25, 96.2% | 0.9607 | - |
| Education (years) | 16.0 ± 2.3 | 15.9 ± 2.7 | 15.5 ± 2.6 | 0.6470 | - |
| BMI | 27.3 ± 4.7 | 27.1 ± 4.6 | 26.1 ± 4.3 | 0.4780 | - |
| **Clinical Parameters** |  |  |  |  |  |
| Family history of PD<br>(yes, %) | 10, 10.6% | 19, 18.1% | 3, 11.5% | 0.6806 | - |
| Duration (years) | 0.6 ± 0.6 | 0.6 ± 0.5 | 0.6 ± 0.6 | 0.8829 | - |
| LEDD (mg/day) | 0 | 0 | 0 | - | - |
| MDS-UPDRS-part I | 5.6 ± 4.0 | 5.9 ± 3.9 | 5.0 ± 3.1 | 0.5969 | - |
| MDS-UPDRS-part II | 5.4 ± 3.8 | 6.2 ± 4.3 | 6.3 ± 4.7 | 0.3429 | - |
| MDS-UPDRS-part III<br>in OFF state | 20.5 ± 8.5 | 22.8 ± 9.7 | 20.2 ± 9.2 | 0.1675 | - |
| MDS-UPDRS-part IV | 0 | 0 | 0 | - | - |
| MDS-UPDRS total score | 31.5 ± 12.2 | 34.8 ± 13.7 | 31.5 ± 14.2 | 0.1607 | - |
| HY in OFF state | 1.5 ± 0.5 | 1.6 ± 0.5 | 1.4 ± 0.5 | 0.3519 | - |
| MoCA | 27.2 ± 2.4 | 27.0 ± 2.4 | 27.1 ± 2.4 | 0.8830 | - |
| SCOPA-AUT | 9.1 ± 6.9 | 9.7 ± 5.6 | 10.6 ± 7.7 | 0.5235 | - |
| RBDSQ | 3.6 ± 2.5 | 4.3 ± 2.9 | 4.3 ± 2.6 | 0.2035 | - |
| UPSIT | 22.3 ± 7.7 | 21.1 ± 7.8 | 19.9 ± 8.2 | 0.3486 | - |
| STAI | 64.7 ± 17.9 | 67.1 ± 18.2 | 59.5 ± 17.9 | 0.1447 | - |
| PIGD<br>phenotype (yes, %) | 24, 25.5% | 28, 26.7% | 3, 11.5% | 0.2611 | - |
| Diffuse Malignant<br>phenotype (yes, %) | 10, 10.9% | 13, 12.4% | 4, 16.0% | 0.2349 | - |

### Clinical Trajectories of Cluster-Derived Biological Subtypes Over 5 Years

We then assessed longitudinal changes in clinical variables by using mixed analysis of covariance (mixed ANCOVA), adjusted for age and sex, to investigate differences in disease progression across clusters over the 5-year follow-up period. Clinical parameters at 5-year follow-up were available for n=173 subjects (Table 4). No significant differences in the progression of motor or non-motor features were observed between clusters.

**Table 4.**
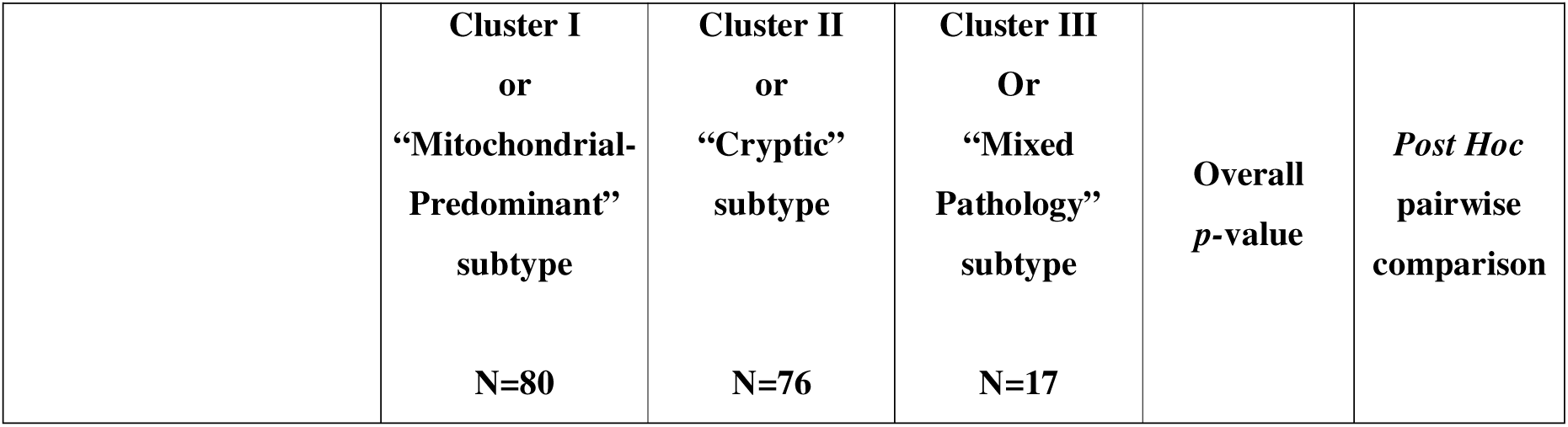
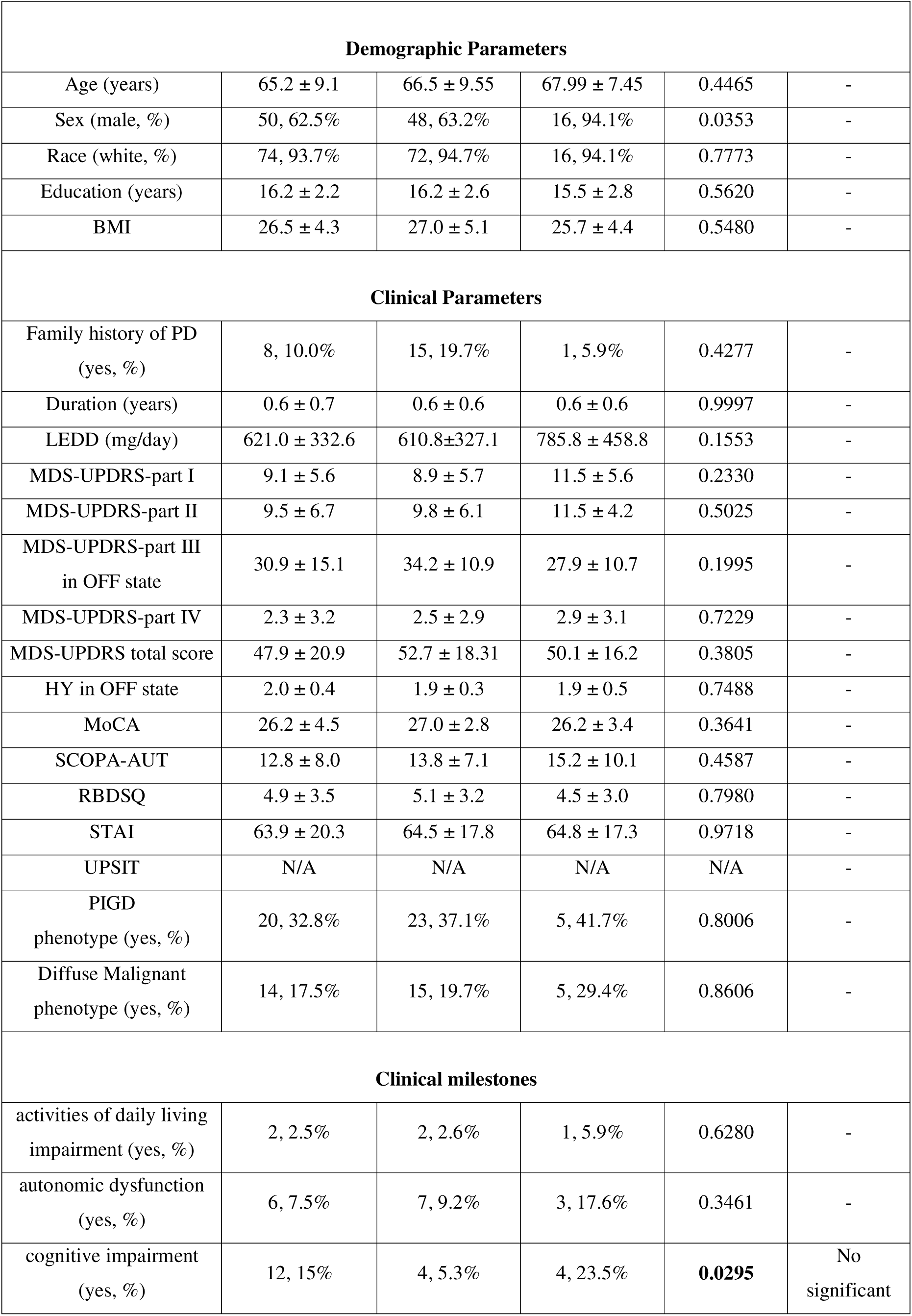
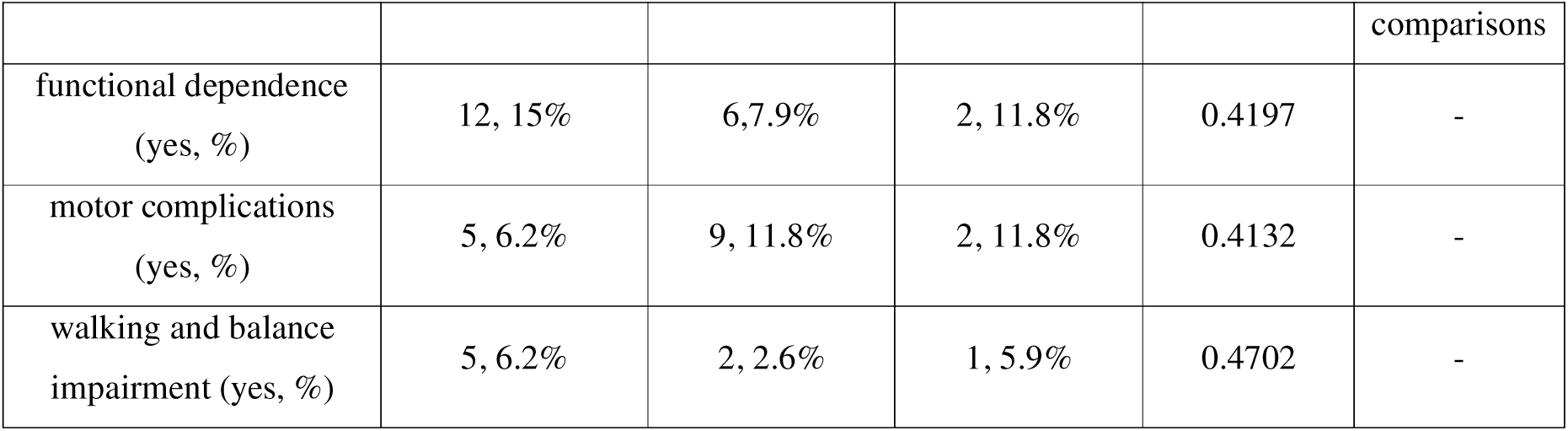
Comparison of demographic and clinical parameters across clusters at 5-year follow-up. Continuous variables are presented as mean ± standard deviation, and categorical variables as number, frequency. For overall *p*-value, statistical significance is marked in bold. For *Post Hoc* pairwise comparisons, only statistically significant comparisons are reported.; Abbreviations: n, number; BMI, body mass index; LEDD, levodopa equivalent daily dose; MDS-UPDRS-part I, II, III, IV and total score, Movement Disorder Society-Unified PD Rating scale part part I, II, III, IV and total score; HY, Hoehn and Yahr scale; MoCA, Montreal cognitive assessment; SCOPA-AUT, Scales for Outcomes in PD-Autonomic dysfunction; RBDSQ, RBD Screening Questionnaire; UPSIT, University of Pennsylvania Smell Identification Test; STAI, State-Trait Anxiety Inventory; PIGD, Postural Instability and Gait Difficulty; N/A, not available.

|  | Cluster I<br>or<br>“Mitochondrial-Predominant”<br>subtype<br>N=80 | Cluster II<br>or<br>“Cryptic”<br>subtype<br>N=76 | Cluster III<br>Or<br>“Mixed<br>Pathology”<br>subtype<br>N=17 | Overall<br>*p*-value | *Post Hoc*<br>pairwise<br>comparison |
| --- | --- | --- | --- | --- | --- |

| Demographic Parameters |  |  |  |  |  |
| --- | --- | --- | --- | --- | --- |
| Age (years) | 65.2 ± 9.1 | 66.5 ± 9.55 | 67.99 ± 7.45 | 0.4465 | - |
| Sex (male, %) | 50, 62.5% | 48, 63.2% | 16, 94.1% | 0.0353 | - |
| Race (white, %) | 74, 93.7% | 72, 94.7% | 16, 94.1% | 0.7773 | - |
| Education (years) | 16.2 ± 2.2 | 16.2 ± 2.6 | 15.5 ± 2.8 | 0.5620 | - |
| BMI | 26.5 ± 4.3 | 27.0 ± 5.1 | 25.7 ± 4.4 | 0.5480 | - |
| Clinical Parameters |  |  |  |  |  |
| Family history of PD<br>(yes, %) | 8, 10.0% | 15, 19.7% | 1, 5.9% | 0.4277 | - |
| Duration (years) | 0.6 ± 0.7 | 0.6 ± 0.6 | 0.6 ± 0.6 | 0.9997 | - |
| LEDD (mg/day) | 621.0 ± 332.6 | 610.8±327.1 | 785.8 ± 458.8 | 0.1553 | - |
| MDS-UPDRS-part I | 9.1 ± 5.6 | 8.9 ± 5.7 | 11.5 ± 5.6 | 0.2330 | - |
| MDS-UPDRS-part II | 9.5 ± 6.7 | 9.8 ± 6.1 | 11.5 ± 4.2 | 0.5025 | - |
| MDS-UPDRS-part III<br>in OFF state | 30.9 ± 15.1 | 34.2 ± 10.9 | 27.9 ± 10.7 | 0.1995 | - |
| MDS-UPDRS-part IV | 2.3 ± 3.2 | 2.5 ± 2.9 | 2.9 ± 3.1 | 0.7229 | - |
| MDS-UPDRS total score | 47.9 ± 20.9 | 52.7 ± 18.31 | 50.1 ± 16.2 | 0.3805 | - |
| HY in OFF state | 2.0 ± 0.4 | 1.9 ± 0.3 | 1.9 ± 0.5 | 0.7488 | - |
| MoCA | 26.2 ± 4.5 | 27.0 ± 2.8 | 26.2 ± 3.4 | 0.3641 | - |
| SCOPA-AUT | 12.8 ± 8.0 | 13.8 ± 7.1 | 15.2 ± 10.1 | 0.4587 | - |
| RBDSQ | 4.9 ± 3.5 | 5.1 ± 3.2 | 4.5 ± 3.0 | 0.7980 | - |
| STAI | 63.9 ± 20.3 | 64.5 ± 17.8 | 64.8 ± 17.3 | 0.9718 | - |
| UPSIT | N/A | N/A | N/A | N/A | - |
| PIGD<br>phenotype (yes, %) | 20, 32.8% | 23, 37.1% | 5, 41.7% | 0.8006 | - |
| Diffuse Malignant<br>phenotype (yes, %) | 14, 17.5% | 15, 19.7% | 5, 29.4% | 0.8606 | - |
| Clinical milestones |  |  |  |  |  |
| activities of daily living<br>impairment (yes, %) | 2, 2.5% | 2, 2.6% | 1, 5.9% | 0.6280 | - |
| autonomic dysfunction<br>(yes, %) | 6, 7.5% | 7, 9.2% | 3, 17.6% | 0.3461 | - |
| cognitive impairment<br>(yes, %) | 12, 15% | 4, 5.3% | 4, 23.5% | **0.0295** | No<br>significant |

**Table 4. Comparison of demographic and clinical parameters across clusters at 5-year follow-up**
|  |  |  |  |  | comparisons |
| --- | --- | --- | --- | --- | --- |
| functional dependence<br>(yes, %) | 12, 15% | 6, 7.9% | 2, 11.8% | 0.4197 | - |
| motor complications<br>(yes, %) | 5, 6.2% | 9, 11.8% | 2, 11.8% | 0.4132 | - |
| walking and balance<br>impairment (yes, %) | 5, 6.2% | 2, 2.6% | 1, 5.9% | 0.4702 | - |

Clinical progression was further explored by comparing the frequency of clinical milestones, as defined according to PPMI criteria, at the 5-year follow-up, including activities of daily living impairment, autonomic dysfunction, cognitive impairment, functional dependence, motor complications, and walking and balance impairment^41^. Fisher’s exact test revealed an overall difference in the frequency of subjects reaching the cognitive impairment milestone across clusters; however, no statistically significant pairwise comparisons were identified in the post hoc analysis. No differences were observed for the remaining clinical milestones.

Eventually, the distribution of clinical subtypes, determined at 5-year follow-up using the TD/PIGD and MMP/IM/DM classifications, was similar across cluster-derived biological subtypes (Table 4, Figure 4).

## Discussion

The complexity and heterogeneity of iPD pathophysiology suggest the existence of distinct biological subtypes, the identification of which remains a top priority in the field of PD research^23,42^. Using a data-driven approach based on a set of 22 fluid biomarkers measured in 225 subjects with *de novo* iPD from the PPMI cohort we identified three subtypes of iPD with presumably different underlying biology. Notably, despite their distinct biomarker signatures, these subtypes exhibited similar clinical presentations at baseline and comparable clinical progression over the mid-term follow-up.

### From Top-Down to Bottom-Up Data-Driven Framework

The present study reverted the paradigm of previous data-driven stratification studies by adopting a “bottom-up” rather than a “top-down” approach^20^. In most data-driven studies, indeed, a so-called “top-down” approach has been used, whereby iPD subtypes were identified using clinical variables, with single or small sets of biomarkers subsequently examined to elucidate their underlying biological basis^3,23^. Although these clinically derived subtypes have proven value for predicting disease course, they have generally provided limited insight into the biological mechanisms underlying iPD heterogeneity^23^. In the present study, we adopted the opposite strategy by using a data-driven “bottom-up” approach. Specifically, cluster analysis was performed exclusively on fluid biomarkers, using a large panel covering the major pathophysiological pathways implicated in iPD, while clinical variables were considered only afterward as post hoc analysis to characterize the clinical presentation and longitudinal progression of the resulting biologically defined subtypes. The “bottom-up” approach adopted here allowed us to focus strictly on the biological heterogeneity existing among subjects with iPD, providing valuable information in the context of precision medicine.

### Biological Interpretation of the Identified Subtypes

The statistical analysis identified three clusters: Cluster I (“Mitochondrial-Predominant” subtype), Cluster II (“Cryptic” subtype), which together included around 90% of subjects, and Cluster III (“Mixed Pathology” subtype), comprising the remaining 10%.

Only participants with a positive CSF-αSyn-SAA outcome were included in the present study based on the study design. Accordingly, α-synucleinopathy was a shared feature across all identified subtypes. The biomarker profile of the “Mitochondrial-Predominant” subtype indicated the relative accumulation of damaged mitochondrial genomes, consistent with increased mitochondrial stress and injury, as the predominant biological trait of this subtype^27–30^. This pathological feature was also observed in the “Mixed Pathology” subtype, where it was accompanied by a broader biomarker profile reflecting a relatively more severe tau pathology, neuroaxonal injury, astrocytic and microglial activation.^36–38^ Notably, the “Mixed Pathology” subtype also exhibited different plasma sphingolipid levels and lower whole-blood GCase activity - although the latter difference only showed a trend toward statistical significance - compared to the other subtypes, suggesting a relative impairment of the lysosomal system^31–33,40^. This finding, however, should be interpreted with caution, as conflicting evidence has been reported regarding the interpretation of sphingolipid metabolism biomarkers in PD^43,44^. Eventually, a “Cryptic” subtype was identified, comprising subjects lacking a characteristic biomarker profile. This subgroup showed slightly different CSF-αSyn-SAA amplification parameters compared with the other two subtypes. The shorter TTT detected in the “Cryptic” subtype theoretically corresponds to a faster accumulation of pathological α-synuclein in the CNS. However, this finding was not considered by the authors as sufficient to label this subtype differently, given the ambiguous literature regarding the quantitative interpretation of CSF-αSyn-SAA and the relatively modest difference in TTT observed across clusters^7,9,11,35,45,46^. It remains to be determined whether the biological pathways underlying the “Cryptic” subtype markedly differ from those involved in the other two subtypes or instead are similar but not adequately captured by the biomarker panel used in the present study. At present, the “Cryptic” subtype appears biologically enigmatic.

### Sex and Age Differences between Biological Subtypes

The three biological subtypes identified in the present study differed in terms of age and sex. Specifically, subjects belonging to the “Mixed Pathology” subtype were predominantly male compared to those from other two subtypes, and approximately six years older than those from the “Mitochondrial Predominant” subtype. In other words, our findings suggest that a more complex pathophysiology is associated with individuals exhibiting greater biological vulnerability, here reflected by older age and male sex^2,38,47^. Indeed, both demographic features have been associated with increased susceptibility to cellular stress in PD and are recognized as independent risk factors for the disease^2,38,47^. Looking at the other side of the coin, our study showed that relatively younger individuals with iPD may exhibit predominant involvement of the mitochondrial system. This highlights the role of mitochondrial dysfunction as one of the earliest and most relevant pathogenic processes in iPD^48–50^. Interestingly, sex differences were not statistically significant in the “Mitochondrial-Predominant” subtype - nor in the “Cryptic” subtype. A similar observation has been reported in monogenic forms of PD primarily characterized by mitochondrial dysfunction, such as *PRKN*-PD, and *PINK1*-PD, where a clear male predominance has not been observed^51,52^. Although speculative, this suggests that the predominant involvement of mitochondrial pathways is not associated with sex differences in either idiopathic or monogenic PD.

### Similarity in Clinical Presentation and Progression across Biological Subtypes

According to our results, the biological heterogeneity of these subtypes was not paralleled by differences in terms of clinical presentation as well as disease progression. Indeed, the three identified subtypes were characterized by a similar burden of motor and non-motor symptoms at both baseline and mid-term follow-up, including when symptoms were profiled through validated clinical subtypes (i.e., TD vs PIGD, or MMP vs IM vs DM). Unlike genetic forms of PD, in which distinct pathogenic variants produce different biological alterations and often distinct clinical phenotypes, these findings indicate that, in iPD, biology and clinical phenotype are not necessarily closely aligned. This is perhaps not surprising. A similar clinical phenotype, indeed, primarily reflects alterations in similar anatomical regions and neuronal circuits, which, however, may be affected by distinct pathophysiological processes. Moreover, inter-individual differences in compensatory and resilience mechanisms also contribute to a non-linear relationship between biological processes and clinical expression^53–55^. The lack of a full correspondence between clinical presentation and underlying biology is an increasingly recognized concept in other movement disorders, such as Primary Familial Brain Calcification (PFBC), where around one-third of patients remain clinically asymptomatic during their lifetime, despite exhibiting brain calcification and carrying pathogenic variants in PFBC genes^56^. A similar phenomenon has also been described in other areas of neurology, including multiple sclerosis (MS) and AD^57–61^. In MS, for example, the term “clinical-radiological paradox” was coined to describe the discrepancy between radiological measures of disease burden and clinical disability, whereas the revised research criteria for AD explicitly distinguish disease biology from clinical stage^57–61^. Recognizing that clinical phenotype and underlying biology are not necessarily closely correlated may facilitate a more unbiased understanding of iPD and support the design of future biology-driven clinical trials.

### Limitation and Strengths

Some limitations must be acknowledged. First, our study lacks a healthy control (HC) group. While the literature reports differences between PD and HCs for most of the biomarkers included in this study, the evidence is less clear for others ^7–9,11–13,27–34,36–40,43,44,46^. Accordingly, the biomarker signature of each cluster reflects the predominance of certain pathological pathways over others, rather than their absolute presence or absence. Second, as reported above, iPD subjects testing negative on CSF-αSyn-SAA were not included due to statistical constraints. Accordingly, pathological mechanisms occurring in subjects without evidence of central synucleinopathy were not investigated. Third, according to the statistical analysis, only n=26 subjects belonged to Cluster III, which slightly reduced the statistical power of the post-hoc comparisons. Fourth, data on environmental risk factors were not available for inclusion in this study. This prevented us from investigating potential associations between exposure to specific toxic agents and biomarker signatures. Eventually, although this study aimed to assess the pathogenic pathways potentially involved in PD pathogenesis as comprehensively as possible, it was intrinsically limited by the availability of biomarkers. Accordingly, several pathological processes were not represented in the analysis, because validated fluid biomarkers are currently lacking or were not available in the PPMI cohort.

Our work has several strengths as well. First, this study was based on a cluster analysis, which allowed us to manage a large dataset from a well-established international cohort using an unbiased approach, compared to hypothesis-driven studies. Importantly, as previously discussed, in this work the classic paradigm of data-driven studies was reversed, giving priority to biomarkers over clinical symptoms, in order to provide a more biology-driven framework for investigating PD pathophysiology. Second, despite some intrinsic limitations discussed above, the selection process led to the inclusion of a wide panel of fluid biomarkers. This panel covered the main pathological pathways putatively involved in the pathogenesis of PD, allowing an extensive investigation of PD biology. Importantly, by integrating biomarkers beyond α-synuclein, this study contributed to expanding the biological framework of PD beyond α-synucleinopathy alone, which might be particularly relevant in light of the recent failure of immune-based α-synuclein trials in PD^62–64^. Above all, our study addressed the main goal of precision medicine, trying to dissect the complexity of PD biological heterogeneity with the hope of facilitating the development of new targeted therapeutic strategies.

## Conclusion

Our study revealed three biological subtypes of iPD characterized by distinct biomarker signatures, suggestive of different underlying biology, but with an overlapping non-distinctive clinical presentation and disease course. Future studies including a broader set of biomarkers are needed to confirm and expand these findings.

## Acknowledgements

Dr. Kang is supported by NIH (R01 NS131658, R01 NS133742, U01 NS113851, U01 NS122419, U01 AG098785, U01 NS113851, RF1 NS126406, OT2OD038130) and Michael J Fox Foundation for Parkinson Research.

Dr. Riboldi is supported by grants from Michael J Fox Foundation, Parkinson’s Foundation, NIH (R01 NS116006; R01 NS133742, OT2OD038130), and received a previous research grant from Prevail Therapeutics.

Dr. Grillo is supported by the Marlene and Paolo Fresco Institute Post-Doctoral Clinical Fellowship.

Dr. Fereshtehnejad is supported by a grant from Parkinson Canada.

Data used in the preparation of this article was obtained on 2025-06-01 from the Parkinson’s Progression Markers Initiative (PPMI) database (www.ppmi-info.org/access-dataspecimens/download-data), RRID:SCR_006431. For up-to-date information on the study, visit www.ppmi-info.org.

PPMI – a public-private partnership – is funded by the Michael J. Fox Foundation for Parkinson’s Research and funding partners, including 4D Pharma, Abbvie, AcureX, Allergan, Amathus Therapeutics, Aligning Science Across Parkinson’s, AskBio, Avid Radiopharmaceuticals, BIAL, BioArctic, Biogen, Biohaven, BioLegend, BlueRock Therapeutics, Bristol-Myers Squibb, Calico Labs, Capsida Biotherapeutics, Celgene, Cerevel Therapeutics, Coave Therapeutics, DaCapo Brainscience, Denali, Edmond J. Safra Foundation, Eli Lilly, Gain Therapeutics, GE HealthCare, Genentech, GSK, Golub Capital, Handl Therapeutics, Insitro, Jazz Pharmaceuticals, Johnson & Johnson Innovative Medicine, Lundbeck, Merck, Meso Scale Discovery, Mission Therapeutics, Neurocrine Biosciences, Neuron23, Neuropore, Pfizer, Piramal, Prevail Therapeutics, Roche, Sanofi, Servier, Sun Pharma Advanced Research Company, Takeda, Teva, UCB, Vanqua Bio, Verily, Voyager Therapeutics, the Weston Family Foundation and Yumanity Therapeutics.

This work is dedicated to the memory of Paolo Fresco, in grateful recognition of his lasting impact on our scientific community.

## Funding

### Funding statement for the preceding 12 months

Dr. Un Kang receives consulting compensation as a SAB member of American Parkinson Disease Association, Amprion, Inc., Nurr-On, and as a consultant for Lundbeck and Ipsen Innovation.

Dr. Giulietta Riboldi: nothing to disclose.

Dr. Piergiorgio Grillo: nothing to disclose.

Dr. Antonio Pisani: nothing to disclose.

Dr. Seyed-Mohammad Fereshtehnejad: nothing to disclose.

## Data availability

Data are available from authors upon reasonable request.

## Supplementary material

Supplementary material is available at Brain online

---

**Supplementary Figure 1.**
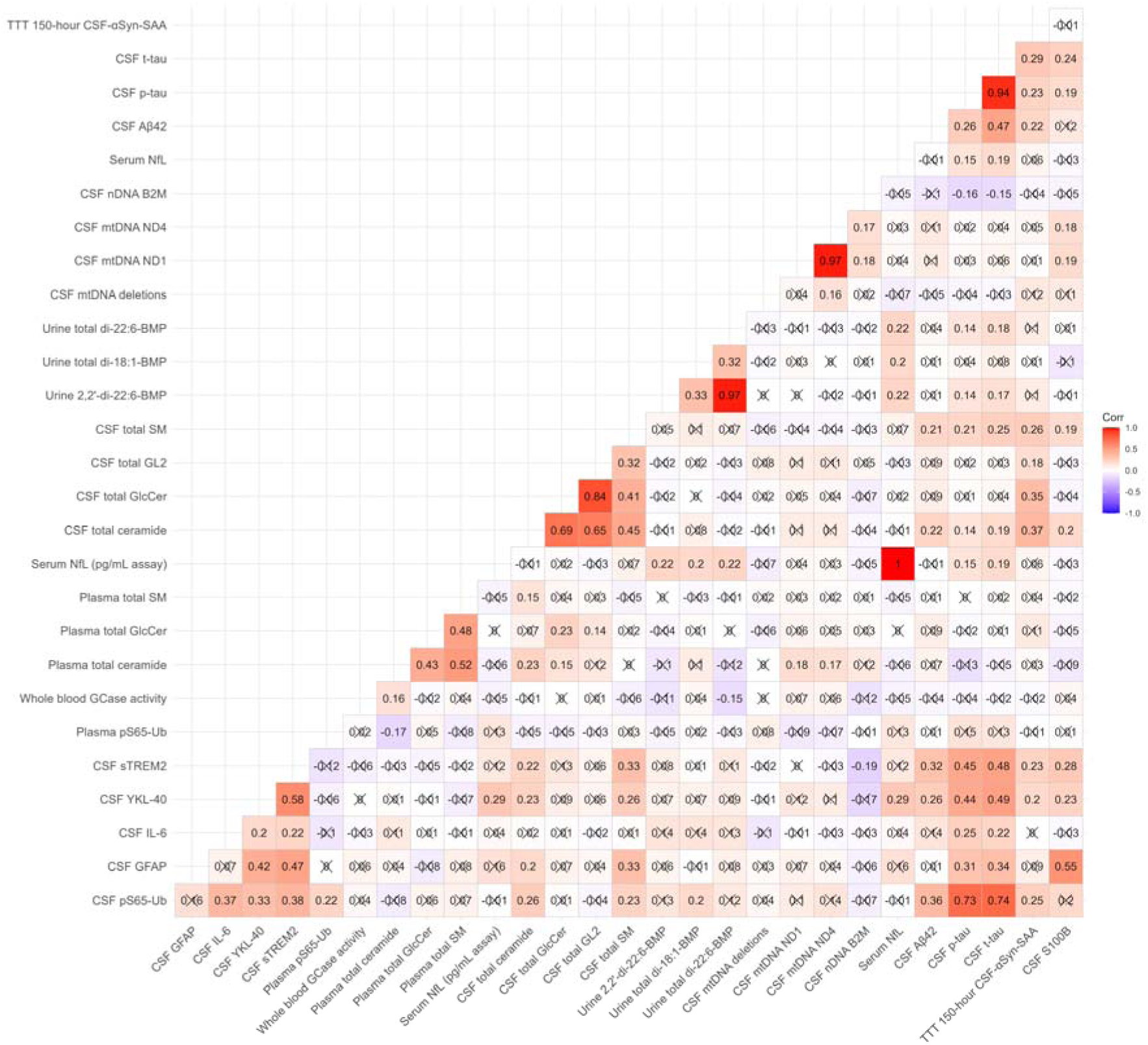
Pairwise correlation matrix of retained fluid biomarkers. Cells show correlation coefficients between biomarker pairs, with color indicating the strength and direction of the correlation. Positive correlations are shown in red and negative correlations in blue, with darker shading indicating stronger correlations. Correlation coefficients are displayed within each cell. Abbreviations: CSF, cerebrospinal fluid; serum NfL, serum neurofilament light chain; CSF Aβ42, CSF amyloid-beta 42; CSF p-tau, CSF phosphorylated tau; TTT from 150-hour CSF-αSyn-SAA, Time to Threshold from 150-hour cerebrospinal fluid α-synuclein seed amplification assay; CSF GFAP, CSF glial fibrillary acidic protein; CSF IL-6, CSF interleukin-6; CSF sTREM2, CSF soluble triggering receptor expressed on myeloid cells 2; CSF mtDNA deletions, CSF mitochondrial DNA deletions; CSF mtDNA ND1, CSF mitochondrial DNA NADH dehydrogenase subunit 1; CSF and plasma pS65-Ub, CSF and plasma phosphorylated serine-65 ubiquitin; whole blood GCase activity, whole blood glucocerebrosidase activity; CSF and plasma total GlcCer, CSF and plasma total glucosylceramide; CSF and plasma total SM, CSF and plasma total sphingomyelin; urine total di-18:1-BMPand total di-22:6-BMP, urine total di-18:1- and total di-22:6-bis(monoacylglycerol)phosphate.

**Supplementary Figure 2.**
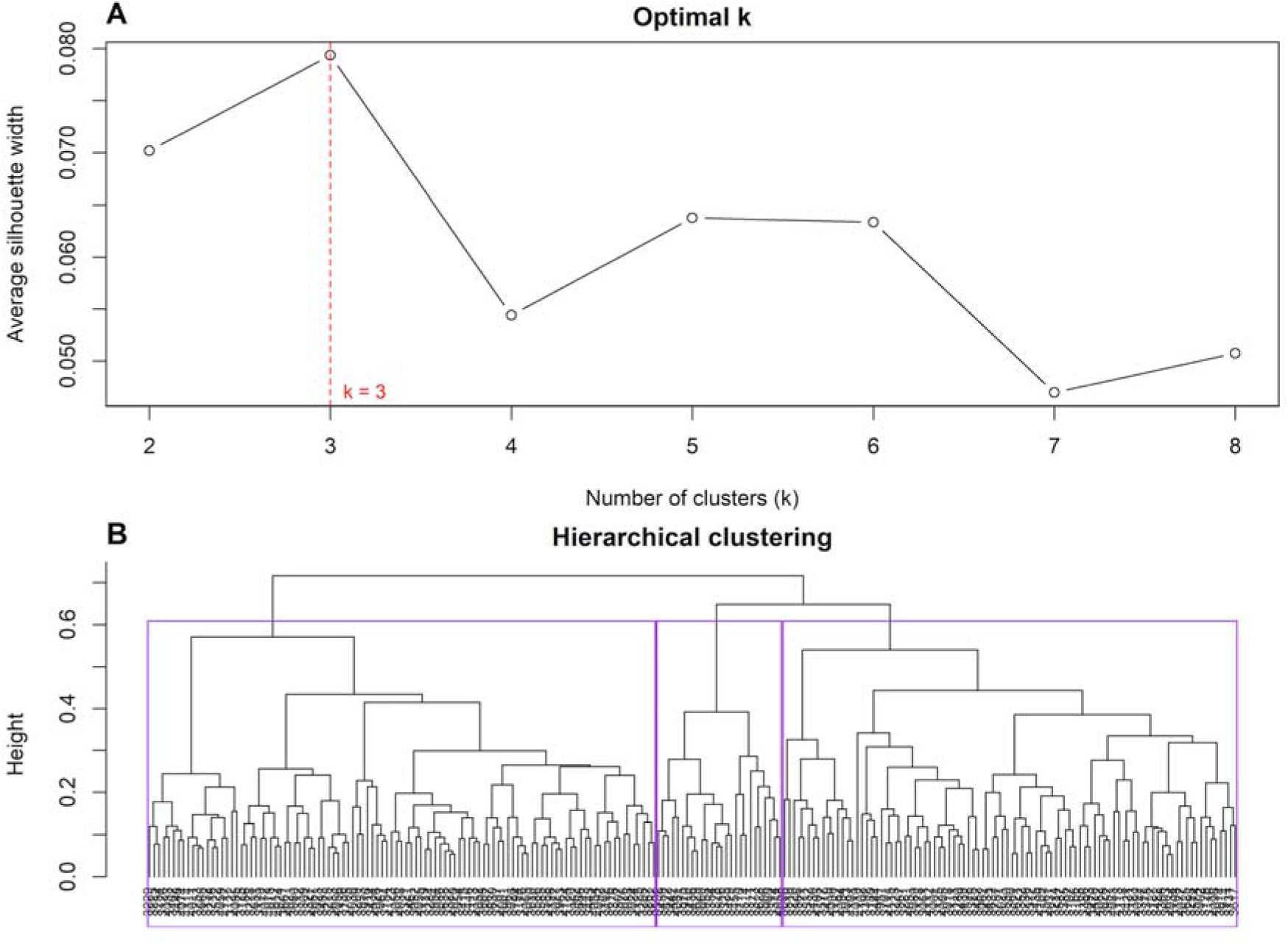
Determination of the optimal number of clusters and hierarchical clustering of biomarker profiles. (A) Average silhouette width across candidate numbers of clusters (k = 2–8). The highest average silhouette width was observed at k = 3, indicating that three clusters provided the optimal partition. The red dashed line marks the selected value of k. (B) Hierarchical clustering dendrogram of study participants based on the biomarkers. The dendrogram was cut to define three clusters, indicated by the colored boxes.

**Supplementary Figure 3.**
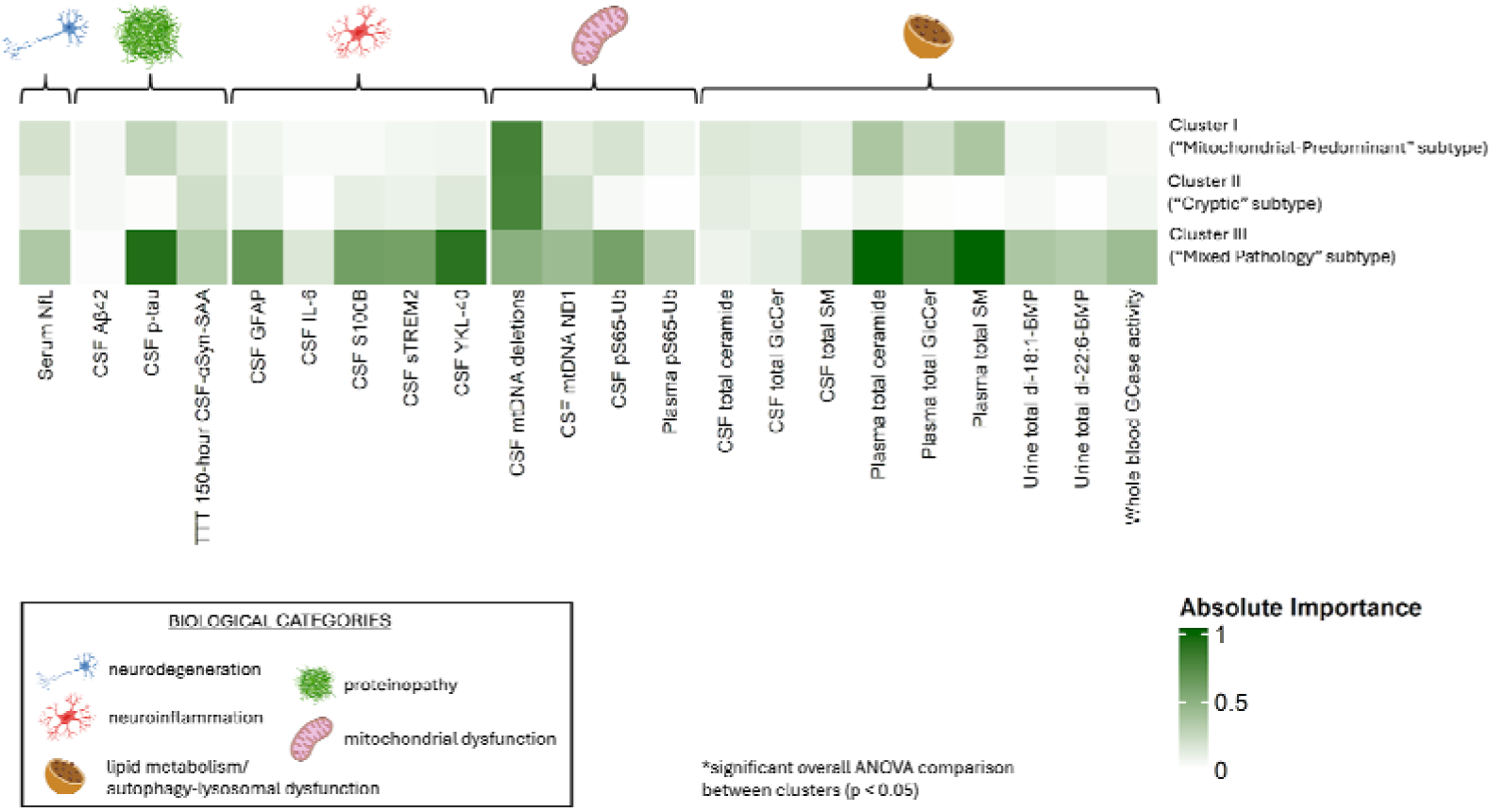
Heatmap of normalized variable importance across clusters. Importance was defined as the absolute value of the standardized effect for each feature and therefore reflects the strength, but not the direction, of deviation from the overall dataset mean. Larger values indicate that a variable more strongly distinguishes a given cluster from the full cohort. Color intensity corresponds to the magnitude of deviation from the global mean, with darker shading indicating greater importance. Variables are grouped by biological category, indicated by distinct icons. Cluster I was renamed as the “Mitochondrial-Predominant” subtype; Cluster II was renamed as the “Cryptic” subtype; Cluster III was renamed as the “Mixed Pathology” subtype *Indicates a statistically significant overall ANOVA comparison between clusters (p < 0.05).

